# Non-inferior survival and enhanced longevity with initial low-dose versus full-dose enzalutamide: a single-centre real-world prostate cancer study

**DOI:** 10.64898/2026.08.28.26361616

**Authors:** Olena Gorobets, Vincent Vinh-Hung

## Abstract

**Background:** Prostate cancer enzalutamide treatment is approved at a standard dose of 160 mg daily. Concerns for real-world patients — older and more fragile than those enrolled in clinical trials — have prompted consideration of initiating treatment with lower doses, but the long-term efficacy of this approach remains unknown. We evaluate the long-term survival and longevity in patients treated with standard versus upfront low-dose enzalutamide.

**Methods:** Retrospective analysis of 151 patients treated with enzalutamide (102 receiving 160 mg; 49 receiving ≤80 mg) between 2014–2021 at the Centre Hospitalier Universitaire de Martinique, with complete follow-up through end of life (98.7% completeness of follow-up). Primary outcomes were overall survival (OS), progression-free survival (PFS), and longevity (attained age).

**Results:** Doses ≤80 mg were associated with longer median OS (36.3 vs. 20.7 months), improved restricted mean OS (difference of 0.7 years, p=0.05), and enhanced longevity (median 82.5 vs. 78.3 years, p=0.004). PSA response rate at 12 weeks was higher with lower-dose (71.4% vs. 48.8%, p=0.016). In multivariable models adjusted for prognostic factors, ≤40 mg compared with 160 mg was non-inferior regarding OS (HR=0.61, 95% CI 0.36–1.06), superior regarding PFS (HR=0.59, 95% CI 0.35–0.99), and superior regarding longevity (HR=0.48, 95% CI 0.28– 0.84). Bone metastasis, poor performance status, PSA response, time to PSA nadir, and disease duration were independent predictors of outcomes. A post-hoc analysis revealed a strong association between dose and physician-prescribing profiles, ranging from “endorse-lowest-dose” to “never-deviate-from-full-dose”.

**Conclusions:** Lower doses of enzalutamide were non-inferior to full-dose. Dose-adapted strategies warrant further investigation.

## Introduction

Enzalutamide is a potent androgen receptor inhibitor. Its value in the treatment of prostate cancer has been demonstrated in several large clinical trials [1, 2]. Initially approved for metastatic castration-resistant prostate cancer, the drug is also effective in hormone-sensitive and in non-metastatic prostate cancer [3]. All trials were conducted using the recommended daily dose of 160 mg. Given the overwhelming evidence of benefit at that dose, there was no reason to prompt a search for a lower dose. The challenge came from real-world observations confronted with poor tolerance of the drug by some patients. Complaints of fatigue and appetite loss could not be ignored. These led to Kimura’s recommendation in 2018 to consider initial dose reduction [4]. The recommendation has seldom been cited; nevertheless, growing evidence steadily showed that a lower dose could be given without loss of antitumor activity [5]. The potential advantage of an upfront reduced dose over the standard dose was prospectively demonstrated in a randomized trial comparing 120 mg with 160 mg enzalutamide in frail patients [6]. The trial found that even a 25% dose reduction induced significantly less fatigue, cognitive side effects, and depressive symptoms, without interfering with efficacy. However, most studies of reduced dose had short follow-up. The long-term efficacy remains unknown. Furthermore, there is no clear indication of which patients would derive the most benefit from a standard or a lower dose of enzalutamide. The present study aims to evaluate long-term survival and longevity using complete follow-up through end of life in prostate cancer patients treated with standard full-dose versus low-dose enzalutamide. A secondary aim is to identify patient or tumor characteristics associated with the survival outcomes.

## Materials and Methods

Records of patients presenting with prostate cancer and receiving enzalutamide therapy between 2014 and 2021 at the urology-oncology division of the Centre Hospitalier Universitaire de Martinique (CHUM) were retrospectively reviewed. No new data were acquired; consent to the retrospective data analyses was waived by the CHUM Institutional Review Board.

The data abstracted were: patient’s age, PSA, Gleason score, and therapy at initial diagnosis, duration of disease from diagnosis to start of enzalutamide, enzalutamide dose prescribed at start and actual dose received during follow-up, patient’s weight, ECOG performance status, pain score on an analog scale of 0 to 10, symptoms and comorbidities, metastatic status, disease localizations, testosterone level, PSA values over time, and indicators of disease progression. Total exposure to enzalutamide was computed for each patient as the cumulative sum of all daily doses actually received. The as-treated average daily dose was computed as the total exposure divided by the patient’s duration of follow-up.

Time was measured in days since starting enzalutamide (Day 0). The baseline PSA was defined as the PSA value estimated on Day 0. It was computed by linear extrapolation from the two most recent PSA measurements preceding Day 0 if available, by forward filling if only one value was available, and was set as fully missing if there were no prior measurements. The PSA at Day 84 (12 weeks, 3 months) was computed by linear interpolation between PSA values obtained before and after 84 days. PSA response was defined as a decrease of ≥50% at 12 weeks. The time to 50% PSA decline was computed by linear interpolation between Day 0 and subsequent measurements. PSA progression was defined as an increase of ≥25% and ≥2 ng/mL from the nadir, or from start of enzalutamide if there was no PSA response [7].

Patients were compared according to the initial enzalutamide dose prescribed: upfront 160 mg (full-dose) vs. upfront ≤80 mg (low-dose). Comparisons used the Student t-test for continuous data and the chi-square test for categorical data. The comparison of survival data used the difference of restricted mean survival time (RMST) or attained age [8], and the logrank test [9, 10]. The event for overall survival (OS) analysis was defined as death from any cause. The event for PSA progression-free survival (PFS) analysis was defined as the first occurrence of either PSA progression or death. The event for longevity (LNG) analysis was the same as for OS, death from any cause, but the LNG analysis, using age as the time scale, estimated the life expectancy from birth [11]. The OS, PFS and LNG time-to-event analyses used the Kaplan-Meier method [12]. Univariable and multivariable Cox regression analyses were conducted to evaluate the association of variables with OS, PFS and LNG [13]. The variables were ranked using the D-measure of prognostic separation [14].

For non-inferiority analysis, the multivariable-adjusted effect of the low dose levels was evaluated using a hazard ratio margin of 1.2, i.e., the median of the margins reported in a meta-epidemiological review [15]. The multivariable adjustments using Cox regression were performed for OS, PFS and LNG outcomes.

In a post-hoc analysis conducted to identify potential selection bias, patterns of dose prescription were examined according to anonymized individual physicians who initiated enzalutamide treatment in the present patient population.

All statistical computations used R version 4.3.2 [16]. The R package ***survival*** was used to compute the Kaplan-Meier survival estimates, the D-measure and the logrank test. Package ***survRM2*** was used for the RMSTs.

## Results

A total of 151 patient records were retrieved. Enzalutamide dose prescription was 160 mg in 102 patients, which included 1 case initially prescribed 120 mg and changed to 160 mg at 3 months, and 11 cases in whom the dose was not explicitly recorded. In accordance with standard institutional practice, the hospital pharmacist contacted the prescribing physician whenever a non-standard dose was specified; conversely, the absence of a dose notation indicated that the standard 160 mg dose had been dispensed as written in the drug monograph. These 11 cases are therefore included in the 160 mg group. The dose was ≤80 mg in 49 patients: 18 patients received 80 mg, 29 received 40 mg, 1 received 40 mg every other day (averaging 20 mg/day), and 1 received 40 mg three days per week (averaging 17.14 mg/day); in total, 31 patients were treated with ≤40 mg/day. Enzalutamide was prescribed for documented metastatic disease in all except 9 patients, of whom 4 had pelvic lymph nodes only (non-metastatic regional recurrence), 2 had prostate local recurrence only, and 3 had undocumented disease extent at the time of enzalutamide.

At last data collection on May 2, 2025, 134 patients had died and 17 were censored. The median follow-up of the censored patients estimated with the reverse Kaplan-Meier method [17] was 2292 days (= 6.28 years). The completeness of follow-up [17] of the study population was 98.7%.

Table 1 summarizes the patients’ characteristics according to the dose group. The characteristics were comparable between the groups except on four features. The ≤80 mg group were significantly older than the 160 mg group by 3 years (mean 78.4 vs. 75.3 years, respectively). The percentage of cardiovascular history in the ≤80 mg group was 44.2%, approximately twice the 23.3% in the 160 mg group. The ≤80 mg group also more frequently had multiple comorbidities (65.1%, versus 45.1% in the 160 mg group). Fewer patients in the ≤80 mg group had previously received abiraterone or chemotherapy compared with patients in the 160 mg group. There was a trend toward poorer ECOG performance status in low-dose patients (37.5% ECOG ≥2, vs. 24.2% in full-dose patients, P=0.099). Low-dose patients were more likely to be truly castration-resistant than full-dose patients, displaying a trend toward more frequent castration (95.9% vs. 83.3%, P=0.096) and a trend toward more documented testosterone assessment (51.0% vs. 38.2%, P=0.137).

**Table 1.**
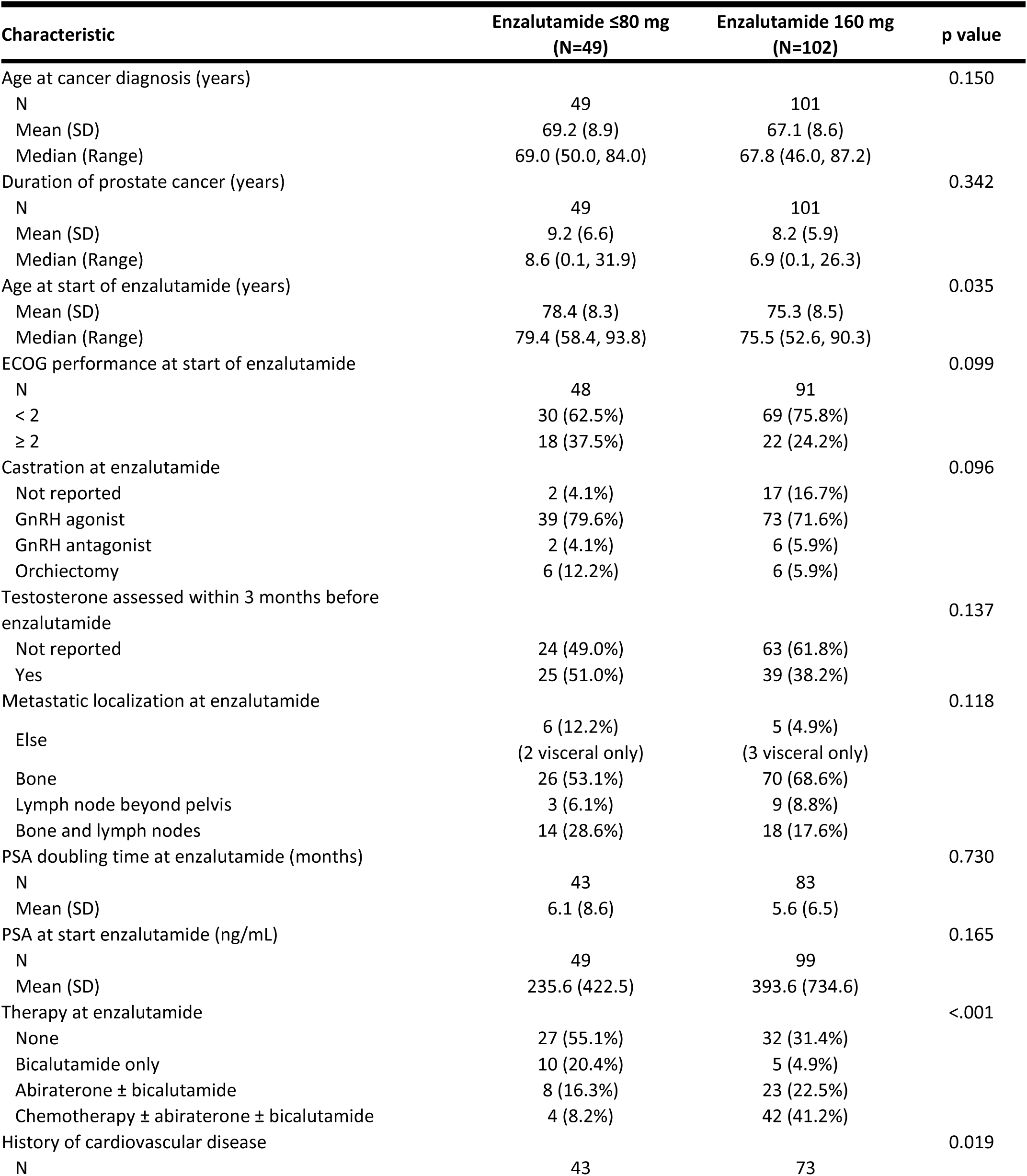
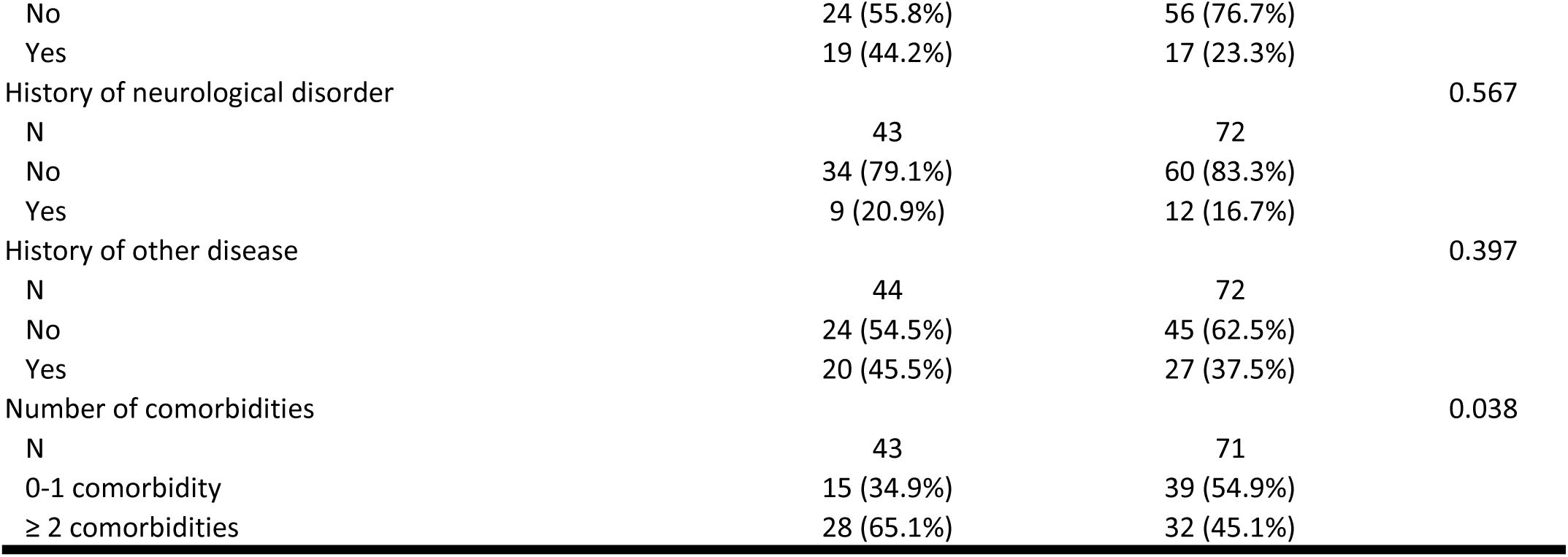
Patients’ main characteristics.

The as-treated exposure was significantly lower in the low-dose group: a mean of 58,404 mg/patient (equivalent to 1,460 capsules of 40 mg) at an average daily dose of 64.8 mg/day (1.62 capsules), compared with 92,173 mg (2,304 capsules) at an average daily dose of 124 mg/day (3.1 capsules) in the full-dose group. Crude unadjusted outcomes showed for the full-dose group a trend toward shorter overall survival (OS), with a median of 20.7 months vs. 36.3 months in the low-dose group, and a shorter progression-free survival (PFS) with a median of 8.5 months (full-dose) vs. 11.7 months (low-dose) (Table 2).

**Table 2.** Patients’ outcomes.

| Crude outcome | Enzalutamide ≤80 mg<br>(N=49) | Enzalutamide 160 mg<br>(N=102) | p value |
| --- | --- | --- | --- |
| As-treated total exposure to enzalutamide (mg/patient) |  |  | 0.027 |
| N | 48 | 102 |  |
| Mean (SD) | 58404 (54149) | 92173 (98035) |  |
| Median (Range) | 51500 (1200, 203024) | 55927 (2080, 496160) |  |
| As-treated enzalutamide dose (mg/day) |  |  | <0.001 |
| N | 48 | 102 |  |
| Mean (SD) | 64.8 (32.4) | 124.0 (41.5) |  |
| Median (Range) | 58.9 (9.3, 160.0) | 138.4 (17.7, 160.0) |  |
| OS: Overall survival (months) |  |  | 0.170 |
| Events | 42 | 92 |  |
| Median Survival | 36.3 | 20.7 |  |
| Crude longevity: Age at last follow-up (years) |  |  | 0.016 |
| Mean (SD) | 81.4 (8.6) | 77.7 (8.7) |  |
| Median (Range) | 82.5 (60.8, 96.5) | 78.3 (53.3, 93.0) |  |
| PSA response: (decline ≥ 50% at 12 weeks) |  |  | 0.016 |
| N | 42 | 86 |  |
| No | 12 (28.6%) | 44 (51.2%) |  |
| Yes | 30 (71.4%) | 42 (48.8%) |  |
| Time to ≥ 50% PSA decline (weeks) |  |  | 0.062 |
| N | 38 | 72 |  |
| Mean (SD) | 6.9 (5.6) | 9.7 (8.0) |  |
| Median (Range) | 4.9 (1.1, 26.4) | 6.9 (0.8, 38.2) |  |
| PSA progression |  |  | 0.007 |
| No | 21 (42.9%) | 22 (21.6%) |  |
| Yes | 28 (57.1%) | 80 (78.4%) |  |
| Time to PSA nadir within 1<sup>st</sup> year (months) |  |  | 0.895 |
| N | 44 | 95 |  |
| Mean (SD) | 5.5 (3.2) | 5.4 (3.3) |  |
| Median (Range) | 5.0 (0.2, 11.5) | 4.6 (0.4, 11.9) |  |
| Time to PSA progression estimate (months) |  |  | 0.008 |
| Events | 28 | 80 |  |
| Median Survival | 19.7 | 10.7 |  |
| PFS: PSA progression-free survival (months) |  |  | 0.059 |
| Events | 49 | 102 |  |
| Median Survival | 11.7 | 8.5 |  |
| PSA at 1 year (ng/mL) |  |  | 0.035 |
| Mean (SD) | 175.4 (424.2) | 398.9 (674.1) |  |
| Median (Range) | 6.7 (0.0, 2273) | 71.9 (0.0, 4517) |  |
| PSA at last follow-up (ng/mL) |  |  | 0.015 |
| Mean (SD) | 218.5 (429.0) | 680.0 (1273.2) |  |
| Median (Range) | 30.9 (0.0, 2273) | 249.3 (0.0, 9984) |  |

The OS RMST difference Δ between the two groups was 0.7 years in favor of ≤80 mg, P = 0.05. The PFS RMST difference Δ was 0.4 years, also in favor of ≤80 mg, but did not reach significance by the test on Δ, P = 0.14 (Figure 1). The non-significance of the logrank test for OS despite the large difference between the groups is a known consequence of non-proportional hazards, for which RMST-based tests have been recommended [18] (Figure 1). Conversely, the logrank test for PFS approached significance owing to better proportionality of the hazards (Figure 1). Regarding longevity, patients in the 160 mg group attained a crude median age of 78.3 years, which was significantly shorter by 4.2 years compared with the median age of 82.5 years attained in the ≤80 mg group (Table 2). The difference was highly significant by both the RMST test (P = 0.008) and the logrank test (P = 0.004) (Figure 1).

**Figure 1.**
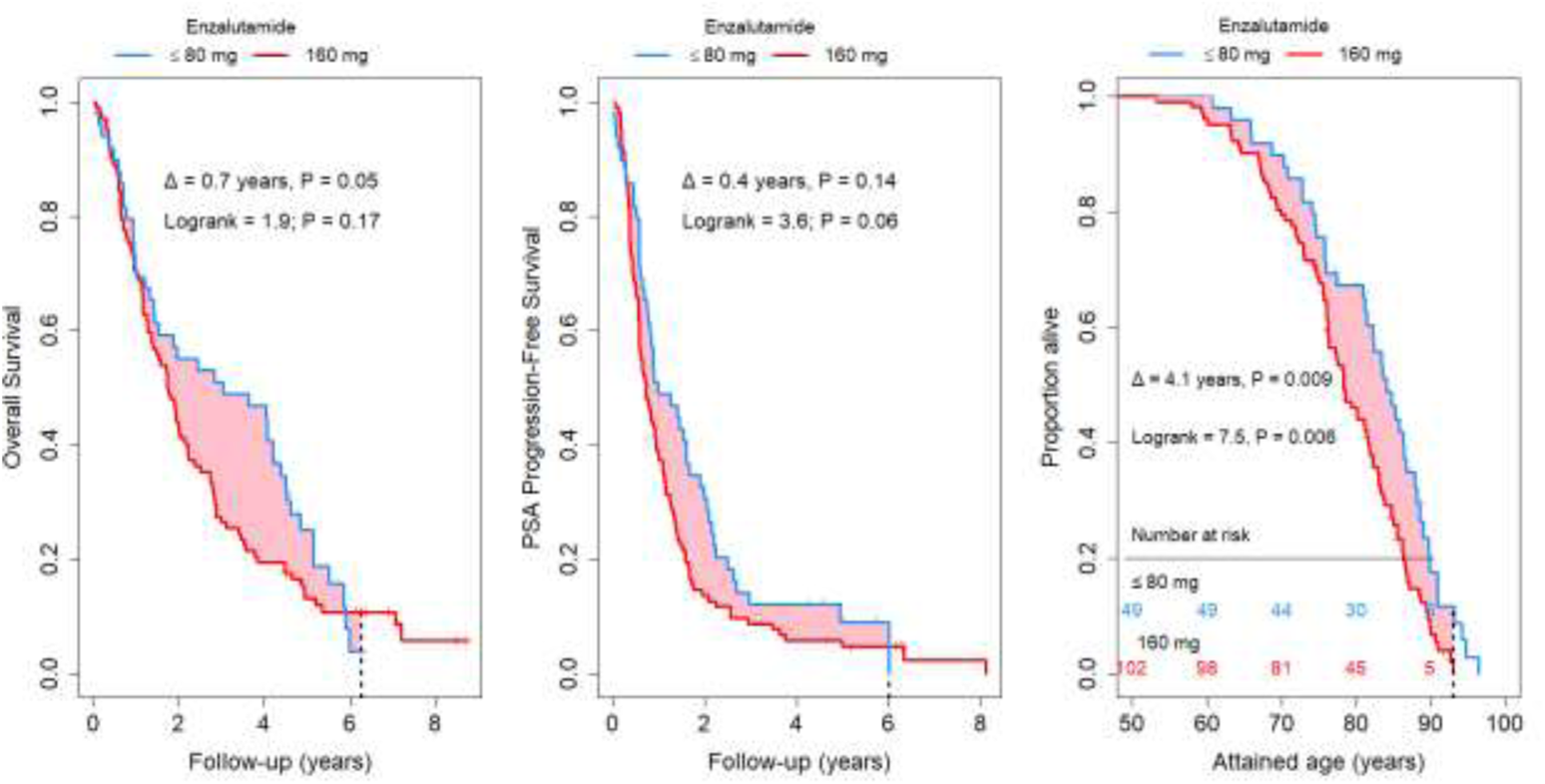
Overall survival, PSA progression free survival and longevity.

Regarding PSA evaluations, the 160 mg group displayed a poorer PSA response at 12 weeks (48.8% vs. 71.4% in the ≤80 mg group) (Figure 2). Other PSA indicators also showed worse outcomes in the 160 mg group, with a shorter median time to PSA progression of 10.7 months vs. 19.7 months in the ≤80 mg group (P = 0.008). At 1-year follow-up, the median PSA was 71.9 ng/mL in the 160 mg group (mean 398.9 ng/mL) vs. 6.7 ng/mL in the ≤80 mg group (mean 175.4 ng/mL), P = 0.035, indicating poorer 1-year PSA control with 160 mg (Table 2).

**Figure 2.**
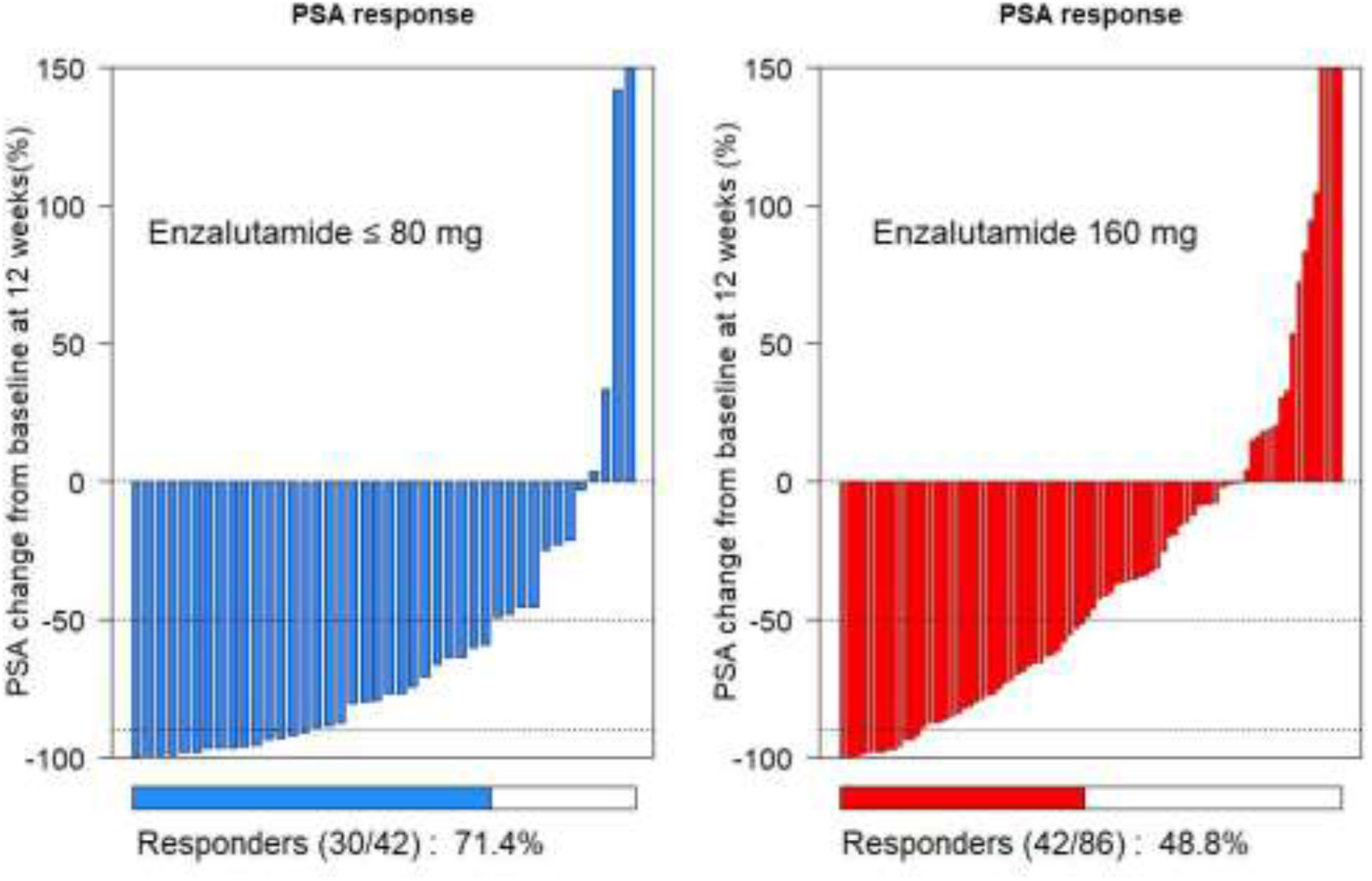
PSA response at 12 weeks.

In a Cox univariable regression analysis regardless of enzalutamide dose, the most important predictors of OS were the various PSA indicators following enzalutamide therapy (Supplemental Table ST1). A D-measure > 0.400 indicates a strong association, corresponding to a hazard ratio near 1.50 (or a 50% relative increased risk of event if the variable was binarized into two equal group sizes). Higher values of PSA at 12 weeks, 9 months, 1 year, or at nadir were predictors of unfavorable OS. A prolonged time to PSA nadir was a favorable predictor. Bone metastases were unfavorable predictors. Poorer ECOG performance status was associated with worse survival. Weight at enzalutamide initiation and time elapsed since prostate cancer diagnosis were significant predictors of overall survival. Predictors of progression-free survival and of longevity were mostly comparable. Of note, the value of PSA at the initiation of enzalutamide was not a significant predictor of survival or longevity. A higher enzalutamide dose was non-significantly associated with worse OS (HR 1.003, indicating a 0.3% relative increase in the risk of death per mg), was significantly associated with worse PFS (P = 0.036), and with worse longevity (P = 0.004) (Supplemental Table ST1).

In a Cox multivariable regression analysis adjusted with categorized dose for overall survival (Table 3), the presence of bone metastasis and poor ECOG score ≥2 were independent predictors of poor OS. PSA response, time to nadir, and a long prostate cancer history preceding enzalutamide were favorable prognostic factors for OS. The categorized dose of 40 mg and 80 mg vs. 160 mg displayed a trend toward poor OS associated with higher doses (Table 3).

**Table 3.** Multivariable Cox regression on overall survival (OS), PSA progression free survival (PFS) and longevity (LNG). HR, hazard ratio. HR > 1 indicates increased risk of event (OS: death, PFS: death or PSA progression, LNG: death) associated with the variable as compared with the reference. HR < 1 indicates decreased risk of event. P, P-value of the HR. The models were computed on N=118 patients, 33 observations were deleted due to missingness. Bottom row: dose modeled as continuous instead of categorized.

| Variable | Reference | HR (OS) | P | HR (PFS) | P | HR (LNG) | P |
| --- | --- | --- | --- | --- | --- | --- | --- |
| Bone metastasis | No bone metastasis | 2.14 | 0.008 | 2.15 | 0.005 | 0.69 | 0.196 |
| ECOG at start of enzalutamide ≥ 2 | ECOG < 2 | 1.63 | 0.035 | 1.46 | 0.098 | 0.70 | 0.126 |
| PSA response: decrease ≥ 50% at 12 weeks | PSA decrease < 50% | 0.34 | 0.000 | 0.35 | 0.000 | 0.86 | 0.463 |
| Time to PSA nadir > 182 days | Time ≤ 182 days | 0.51 | 0.002 | 0.43 | 0.000 | 0.74 | 0.160 |
| Time from prostate cancer diagnosis to enzalutamide > 7 years | Time ≤ 7 years | 0.65 | 0.037 | 0.77 | 0.174 | 0.38 | 0.000 |
| ***Dose categorized:*** |  |  |  |  |  |  |  |
| Enzalutamide 40 mg | Enzalutamide 160 mg | 0.61 | 0.076 | 0.59 | 0.046 | 0.48 | 0.010 |
| Enzalutamide 80 mg | Enzalutamide 160 mg | 1.28 | 0.407 | 1.18 | 0.578 | 0.85 | 0.598 |
| ***Dose continuous:*** |  |  |  |  |  |  |  |
| Enzalutamide (mg) | Per 1 mg increase | 1.003 | 0.203 | 1.003 | 0.101 | 1.005 | 0.015 |

Likewise, the multivariable model of PFS showed qualitatively comparable relationships. In the longevity multivariable analysis (rightmost columns), a strong independent association was found between the lowest dose of enzalutamide and enhanced longevity (P = 0.010) (Table 3). With dose modeled as a continuous variable, each 1 mg increase in enzalutamide dose was associated with a non-significant trend toward worse OS (HR 1.003, P=0.203) and worse PFS (HR 1.003, P=0.101), and with significantly worse longevity (HR 1.005, P=0.015), consistent with the categorical findings. The test of the proportional hazards assumption was satisfied regarding the OS model (P=0.500). The PFS model displayed significant non-proportionality (P=0.005). The LNG global model satisfied proportionality (P=0.525), but ECOG at start of enzalutamide violated proportional hazards (P=0.043).

In the non-inferiority analysis, 95% confidence intervals were applied to assess non-inferiority at a one-sided p-value of 0.025 for the 1.2 margin. The 40 mg dose was non-inferior to 160 mg regarding OS, non-inferior and superior regarding PFS, and both non-inferior and definitively superior to 160 mg regarding longevity (Figure 3). The 80 mg dose compared with 160 mg was inconclusive on all three outcomes (OS, PFS, and longevity).

**Figure 3.**
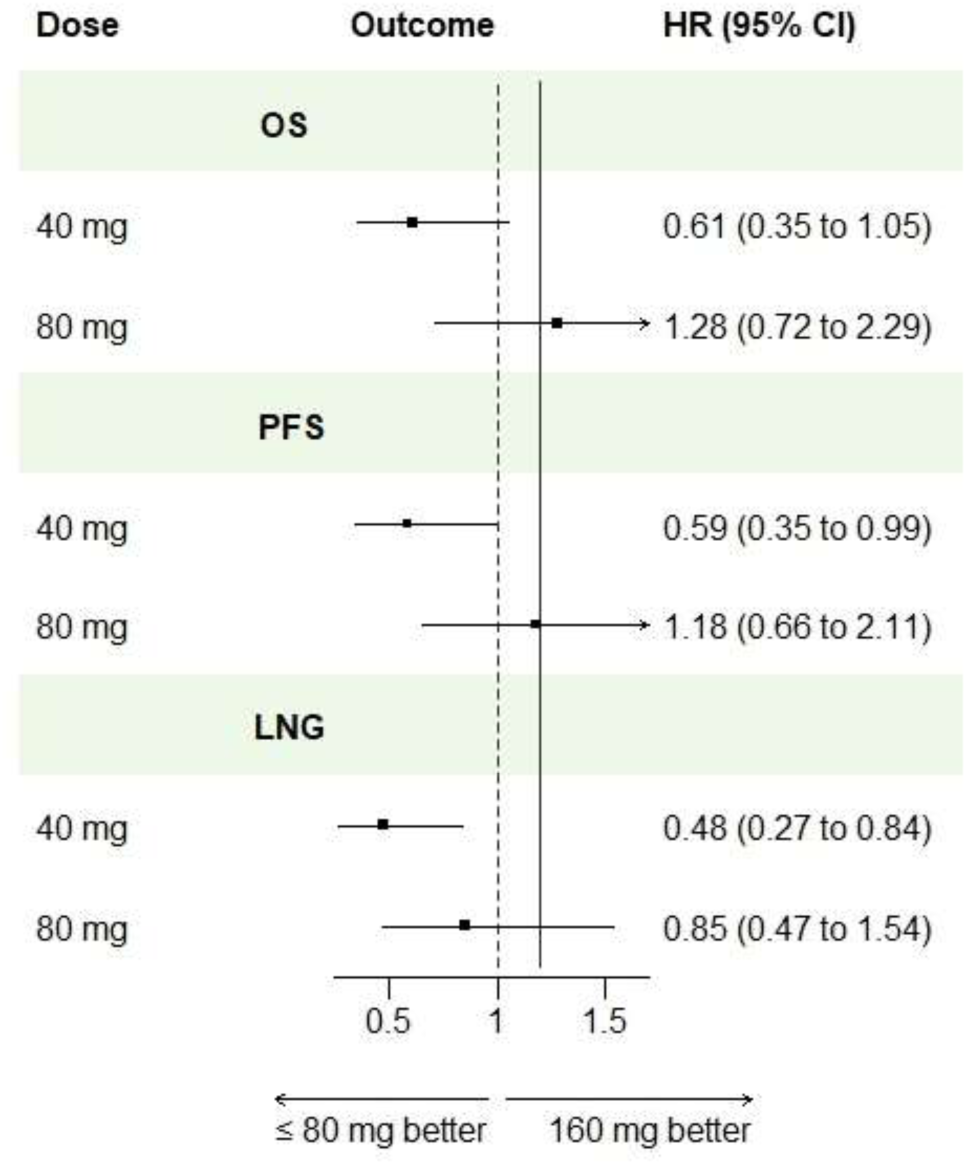
Estimated hazard ratio of dose level for overall survival (OS), PSA-progression free survival (PFS) and longevity (LNG).

In a search of potential selection bias, the enzalutamide doses were tabulated according to the prescribing physicians. A total of 19 physicians were identified. Six physicians accounted for 84% of the 151 initial prescriptions, with 14 to 31 prescriptions per physician. The remaining 16% of the prescriptions were accounted for by 13 physicians, with 1 to 6 prescriptions per physician. Table 4 shows three distinct patterns: 16 of the 19 physicians (84.2%) prescribed predominantly full-dose (Drs A, B, C, and thirteen others – notably, one prescriber never deviated from full-dose), 1 (5.3%) prescribed predominantly 80 mg (Dr D), and 2 (10.5%) prescribed predominantly ≤40 mg (Drs E and F). The association between dose and physician was highly significant (P < 0.001).

**Table 4.** Enzalutamide initial dose, number of cases according to physician (row percent).

| Enzalutamide | ≤20 mg<br>(N=2) | 40 mg<br>(N=29) | 80 mg<br>(N=18) | 120 mg<br>(N=1) | 160 mg<br>(N=101) | All<br>(N=151) | p value |
| --- | --- | --- | --- | --- | --- | --- | --- |
| Physician |  |  |  |  |  |  | <0.001 |
| Dr A | 0 (0%) | 0 (0%) | 0 (0%) | 0 (0%) | **22 (100%)** | 22 |  |
| Dr B | 0 (0%) | 0 (0%) | 1 (5.3%) | 1 (5.3%) | **17 (89.5%)** | 19 |  |
| Dr C | 0 (0%) | 0 (0%) | 3 (9.7%) | 0 (0%) | **28 (90.3%)** | 31 |  |
| Dr D | 0 (0%) | 0 (0%) | **9 (64.3%)** | 0 (0%) | 5 (35.7%) | 14 |  |
| Dr E | 0 (0%) | **16 (80.0%)** | 0 (0%) | 0 (0%) | 4 (20.0%) | 20 |  |
| Dr F | 2 (9.5%) | **12 (57.1%)** | 3 (14.3%) | 0 (0%) | 4 (19.0%) | 24 |  |
| Thirteen others | 0 (0%) | 1 (4.2%) | 2 (8.3%) | 0 (0%) | **21 (87.5%)** | 24 |  |

## Discussion

This retrospective study of 151 prostate cancer patients treated with enzalutamide provides evidence that lower doses (≤80 mg) may offer non-inferior effectiveness with potentially better long-term outcomes compared with the standard 160 mg dose. The reason ≤80 mg was considered low-dose, rather than 120 mg, traces back to the empirical observation of two cases reported by Diakite et al. who discussed the pharmacokinetic rationale [19]. From these cases, some physicians continued with low-dose enzalutamide – not from any sudden epiphany, but from the slow realization that it worked – which led to a personalized approach illustrated in the flowchart (Figure 4).

**Figure 4.**
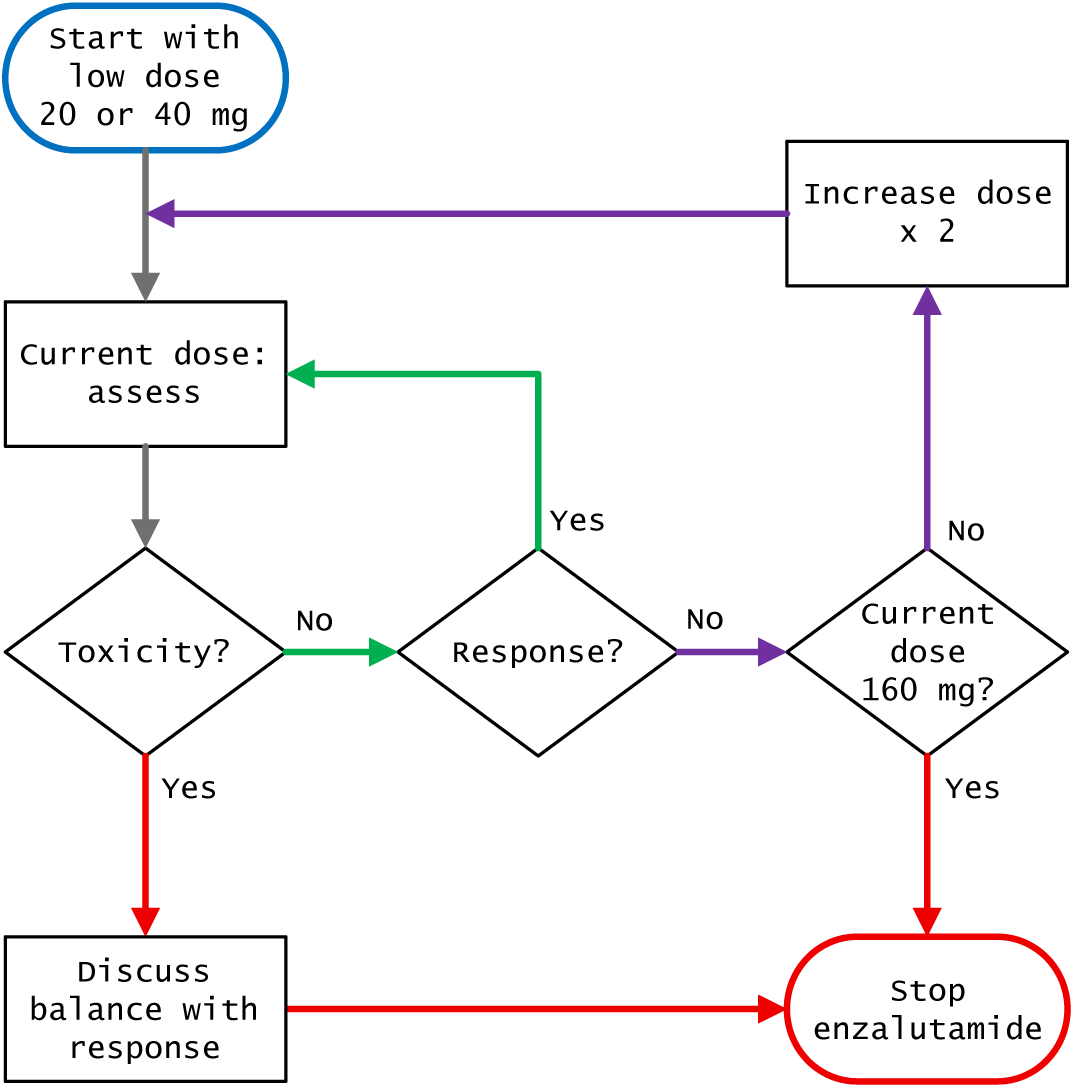
Flowchart of low-dose-adapted enzalutamide. Start with low-dose, maintain the dose unchanged if the surveillance assessment shows response without toxicity (green path). Otherwise consider increasing the dose if no response (purple path), or abandon enzalutamide in case of non-tolerable toxicity.

Our analysis revealed several noteworthy findings. First, patients receiving lower doses demonstrated a trend toward improved survival (median 36.3 vs. 20.7 months). Second, intriguingly, the PSA response rate at 12 weeks was significantly higher in the lower-dose group (71.4% vs 48.8%) (Figure 2). Third, and perhaps most striking, was the significant difference in longevity between the two groups, with lower-dose patients attaining a median age of 82.5 years (despite being older at start and having multiple comorbidities), compared with 78.3 years in the full-dose group. Enhanced longevity was associated with the lowest dose of enzalutamide (40 mg), HR 0.48, P = 0.010 (Table 3). These findings are consistent with drug monitoring studies showing that enzalutamide cohorts were almost universally above pharmacokinetic targets with no demonstrable dose–response relationship [20], suggesting that standard dosing may be excessive. Upfront low-dose enzalutamide may have spared patients from excessive cumulative exposure (Table 2). With complete follow-up, longevity as defined in this study equates to life expectancy from birth. Thus, enhanced longevity most likely reflects not a direct effect of low-dose enzalutamide, but rather a preservation of the patient’s remaining life expectancy.

The multivariable analysis provided insights into factors associated with survival outcomes. Bone metastasis and poor ECOG performance status (≥2) emerged as independent predictors of poor overall survival, consistent with established prognostic factors in advanced prostate cancer [21]. PSA response, longer time to PSA nadir, and longer disease history preceding enzalutamide were favorable prognostic factors [22].

Several limitations merit consideration. The 80 mg dose considered separately was inconclusive across all three outcomes, possibly due to the smaller number of cases (Table 4). Missing values in covariates degraded the efficiency of multivariable analyses. A seven-degree-of-freedom model was the best we could achieve, at the cost of excluding 21.9% of cases (Table 3). Information on adverse events and quality of life was not systematically collected, limiting our ability to assess the impact of dose reduction on toxicity. As a retrospective study, treatment allocation was not randomized. We uncovered a strong relationship between dose and physician (Table 4). Understanding the table requires taking into account the context of the CHUM and its cancer patient general pathway. Patients are referred following a multidisciplinary team (MDT) meeting decision recommending therapy. The CHUM is a public institution. Physicians do not choose their patients. Appointments are assigned according to physicians’ available scheduling slots. By chance alone — analogous to a Galton board experiment in which a bead randomly falls into a bin — a patient was assigned to a physician who happened to be either a full-dose prescriber (regardless of patient condition) or a low-dose prescriber (adapting dose to patient age). Naturally, management differed, such as the trend of more scrutiny of hormone status in low-dose patients (Table 1). It is not the scope of this paper to elucidate how oncologists’ education, training, exposure to pharma representatives, experience, or other factors affected outcomes as demonstrated in other cancer studies [23, 24]. The main point is that a patient receiving any given initial dose was, as far as we could ascertain, the result of an unbiased random process. Inadvertently, this constitutes not a limitation but a strength arising from happenstance.

Further dissecting the issue of selection in light of randomized controlled trials: the PREVAIL double-blind randomized trial compared placebo versus enzalutamide 160 mg in chemotherapy-naive patients presenting with metastatic castration resistant prostate cancer [1]. The enzalutamide arm median OS was 35.3 months; the patients’ median age at inclusion was 72 years, meaning they were approximately 75 years old at the time of death. PREVAIL required an ECOG status of 0–1 for inclusion, and excluded any “co-morbidity that, in the judgment of the Investigator, would make the patient inappropriate for enrollment”, “clinically significant cardiovascular disease”, “history of arrhythmias”, etc. (information buried in the various protocol amendments, which no medical practitioner would ever have time to dig out). Our low-dose patients were older at the start of enzalutamide, with a median age of 78.4 years (Table 1); they had multiple comorbidities, and 37.5% had an ECOG status ≥2, which would have excluded them from any trial inclusion — and yet they survived to approximately age 81.4 years (Table 2). The PREVAIL population and our patients thus differed markedly: our patients were 6.4 years older on average and would largely have been ineligible for PREVAIL.

The AFFIRM double-blind randomized trial compared placebo versus enzalutamide 160 mg in patients with castration-resistant prostate cancer after chemotherapy [2]. The enzalutamide arm median OS was 18.4 months. The patients’ median age was 69 years (AFFIRM supplementary material), meaning AFFIRM patients died at approximately age 70.5 years. AFFIRM required an ECOG status of 0–2; in the reality of the conducted trial, 91.3% of the patients had an ECOG 0–1, 71.8% had a pain score <4, and the baseline PSA was 107.7 ng/mL. AFFIRM exclusion criteria also covered comorbidities, and additionally excluded patients receiving aminophylline, theophylline, risperidone, amiodarone, droperidol, insulin, erythromycin, phenothiazine, imipramine, venlafaxine, and many other drugs. Granted, few of our low-dose patients had received chemotherapy — 8.2% compared with 41.2% of the full-dose patients (Table 1) — and 81.7% had bone metastases (vs. 92.2% in AFFIRM). But by all other measures, our patients were far less fit than the AFFIRM cohort and had a baseline PSA of 235.6 ng/mL, twice as high as the AFFIRM baseline of 107.7 ng/mL. Again, the populations differ. What matters here is that the highly selected AFFIRM population was not representative of our patients (recalling that our patients’ median age was 78.4 years, 9.4 years older than AFFIRM). Thus, transposing dose inferences from AFFIRM to our population appears unwarranted, if only because many drugs commonly used by our patients were excluded from AFFIRM. Indeed, OS in our patients showed a severe rate of death: regardless of dose, 30% died within the first year (Figure 1).

Pending a meta-analysis [25] and a major upcoming clinical trial that will also seek biomarkers to guide dose reduction [26], our findings support consideration of personalized enzalutamide dosing in patients who are older, have poor performance status, or have significant comorbidities. With the caveat that our PFS model displayed non-proportional hazards (Table 3), the non-inferiority of the 40 mg dose regarding overall survival, combined with its putative superior performance regarding progression-free survival and longevity, challenges the one-size-fits-all full-dose approach.

## Conclusion

This retrospective analysis suggests that lower doses of enzalutamide (≤80 mg), and particularly the 40 mg dose, may offer non-inferior or even superior preservation of long-term outcomes compared with the standard 160 mg dose in selected prostate cancer patients. These findings support a personalized approach in which starting with a reduced dose is considered for patients who may benefit from improved tolerability without compromising efficacy. Prospective randomized trials are warranted to confirm these observations and establish optimal patient selection criteria for upfront reduced-dose enzalutamide therapy.

## Data Availability

All data produced in the present study are available upon reasonable request to the authors

## Statements

### Author Contributions

Conceptualization, YJD, VVH; Methodology, OG, VVH; Software, VVH; Validation, OG, YJD; Formal Analysis, VVH; Investigation, OG, YJD, VVH; Resources, VVH; Data Curation, YJD, VVH; Writing – Original Draft Preparation, OG, VVH; Writing – Review & Editing, OG, YJD, VVH; Visualization, OG, VVH; Supervision, VVH; Project Administration, VVH; Funding Acquisition, None.

### Ethics

The study received ethics approval from the Centre Hospitalier Universitaire de Martinique Institutional Review Board. The study was performed in accordance with the Declaration of Helsinki. Patient consent was waived due to no new data being acquired.

## Acknowledgments

In memory to all who passed, whom we will always miss. With heartfelt thanks to the families, and to all colleagues who contributed. The bulk of the study was conducted while Olena Gorobets and Vincent Vinh-Hung were attached to the Centre Hospitalier Universitaire de Martinique. Manuscript editing used Generative Artificial Intelligence: OpenAI ChatGPT for the first draft, thereafter Anthropic Claude to consolidate the structure and to address initial shortcomings.

## Data sharing

Data is available from the corresponding author on sharing request.

