## Supplemental Table ST1 for "Non-inferior survival and enhanced longevity with initial low-dose versus full-dose enzalutamide: a single-centre real-world prostate cancer study"

### Supplemental Material

**Supplemental Table ST1. Cox univariable predictors of overall survival (OS), PSA-progression free survival (PFS), and longevity (LNG).**

For each variable, the row displays the hazard ratio H(xx), its P value P(xx), and the D-measure of prognostic separation D(xx), for the outcome xx (OS, PFS, LNG). Rows ordered by decreasing D(OS).

| Variable (unit) | H(OS) | P(OS) | D(OS) | H(PFS) | P(PFS) | D(PFS) | H(LNG) | P(LNG) | D(LNG) |
|---|---|---|---|---|---|---|---|---|---|
| PSA at 9 months (ng/mL) | 1.001 | 0.000 | 1.489 | 1.001 | 0.000 | 1.583 | 1.000 | 0.005 | 0.593 |
| PSA at 1 year (ng/mL) | 1.001 | 0.000 | 1.472 | 1.001 | 0.000 | 1.499 | 1.000 | 0.002 | 0.620 |
| PSA nadir in year 1 (-% baseline) | 1.003 | 0.000 | 0.966 | 1.004 | 0.000 | 1.271 | 1.004 | 0.000 | 0.505 |
| Time to year 1 PSA nadir (days) | 0.995 | 0.000 | 0.921 | 0.995 | 0.000 | 0.907 | 0.996 | 0.001 | 0.621 |
| PSA at 12 weeks (ng/mL) | 1.006 | 0.000 | 0.920 | 1.009 | 0.000 | 1.141 | 1.005 | 0.000 | 0.471 |
| Bone metastasis (yes/no) | 2.274 | 0.002 | 0.725 | 2.086 | 0.003 | 0.649 | 0.942 | 0.819 | 0.053 |
| PSA at enzalutamide (ng/mL) | 1.0002 | 0.056 | 0.583 | 1.0002 | 0.108 | 0.566 | 0.99999 | 0.934 | -0.094 |
| Weight at enzalutamide (kg) | 0.977 | 0.002 | 0.553 | 0.980 | 0.005 | 0.456 | 1.001 | 0.871 | 0.020 |
| ECOG at enzalutamide (ordinal 0-4) | 1.307 | 0.005 | 0.446 | 1.250 | 0.016 | 0.372 | 0.800 | 0.029 | 0.404 |
| PSA doubling time (days) | 0.999 | 0.052 | 0.432 | 0.999 | 0.028 | 0.358 | 0.999 | 0.207 | 0.346 |
| Years before enzalutamide (years) | 0.961 | 0.012 | 0.401 | 0.974 | 0.081 | 0.298 | 0.940 | 0.000 | 0.567 |
| Prior bicalutamide (yes/no) | 0.701 | 0.111 | 0.324 | 0.758 | 0.195 | 0.254 | 0.908 | 0.667 | 0.088 |
| Cardiovascular history (yes/no) | 1.392 | 0.116 | 0.323 | 1.223 | 0.331 | 0.196 | 0.821 | 0.372 | 0.192 |
| Prior chemotherapy (yes/no) | 1.085 | 0.162 | 0.311 | 1.150 | 0.014 | 0.439 | 1.186 | 0.001 | 0.724 |
| Hypertension (yes/no) | 0.739 | 0.139 | 0.303 | 0.840 | 0.375 | 0.175 | 0.860 | 0.458 | 0.151 |
| Dose enzalutamide (mg) | 1.003 | 0.106 | 0.297 | 1.004 | 0.036 | 0.364 | 1.005 | 0.004 | 0.535 |
| Prior abiraterone (yes/no) | 1.140 | 0.134 | 0.282 | 1.222 | 0.022 | 0.387 | 1.190 | 0.069 | 0.294 |
| Age at cancer diagnosis (years) | 1.017 | 0.110 | 0.246 | 1.004 | 0.675 | 0.084 | 0.926 | 0.000 | 1.121 |
| PSA at cancer diagnosis (ng/mL) | 1.00004 | 0.797 | 0.239 | 1.0001 | 0.632 | 0.341 | 1.0002 | 0.154 | 0.328 |
| Testosterone assessed (yes/no) | 0.926 | 0.643 | 0.204 | 0.826 | 0.263 | 0.341 | 0.959 | 0.819 | 0.136 |
| Gleason score (ordinal 4-10) | 1.088 | 0.268 | 0.152 | 1.003 | 0.967 | 0.001 | 1.197 | 0.014 | 0.345 |
| Lymph node metastasis (yes/no) | 0.865 | 0.448 | 0.140 | 0.849 | 0.372 | 0.158 | 1.041 | 0.834 | 0.039 |
| Weight at cancer diagnosis (kg) | 0.9996 | 0.974 | 0.137 | 1.003 | 0.781 | 0.106 | 1.010 | 0.378 | 0.260 |
| Neurologic comorbidity (yes/no) | 1.071 | 0.788 | 0.063 | 1.058 | 0.819 | 0.051 | 0.770 | 0.308 | 0.238 |
| Lung metastasis (yes/no) | 0.939 | 0.786 | 0.057 | 0.691 | 0.103 | 0.337 | 1.531 | 0.072 | 0.387 |
| Pain at enzalutamide (ordinal 0-10) | 1.006 | 0.842 | 0.051 | 1.008 | 0.797 | 0.061 | 0.979 | 0.505 | 0.146 |
| Diabetes comorbidity (yes/no) | 1.012 | 0.962 | 0.011 | 0.995 | 0.983 | 0.005 | 0.889 | 0.635 | 0.110 |
| Age at enzalutamide (years) | 0.997 | 0.806 | 0.002 | 0.990 | 0.346 | 0.100 | 0.656 | 0.000 | 5.300 |
| Testosterone at enzalutamide (ng/mL) | 1.223 | 0.351 | -0.080 | 1.140 | 0.556 | -0.145 | 0.928 | 0.762 | 0.204 |
